# COVID-19 vaccine uptake among healthcare workers in the fourth country to authorize BNT162b2 during the first month of rollout

**DOI:** 10.1101/2021.01.29.21250749

**Authors:** Mazin Barry, Mohamad-Hani Temsah, Fadi Aljamaan, Basema Saddik, Ayman Al-Eyadhy, Shuliweeh Alenezi, Nurah Alamro, Abdullah N Alhuzaimi, Ali Alhaboob, Khalid Alhasan, Fahad Alsohime, Ali Alaraj, Rabih Halwani, Amr Jamal, Omar Temsah, Fahad Alzamil, Ali Somily, Jaffar A. Al-Tawfiq

## Abstract

**Background:** The Kingdom of Saudi Arabia (KSA) was the fourth country in the world to authorize the BNT162b2 coronavirus disease 2019 (COVID-19) vaccine, which it rolled out on December 17, 2020 and first targeted at healthcare workers (HCWs). This study assesses vaccine uptake among this group during the first month of its availability.

**Methods:** A national cross-sectional, pilot-validated, self-administered survey was conducted among HCWs in the KSA between December 27, 2020 and January 3, 2021. The survey included sociodemographic details, previous contact with COVID-19 patients, previous infection with COVID-19, receiving (or registering with the Ministry of Health website to receive) the COVID-19 vaccine, sources of HCWs’ information on vaccines, awareness of emerging variants of concern, and anxiety level using the 7-item Generalized Anxiety Disorder assessment. A descriptive bivariate analysis and multivariate logistic binary regression analysis were performed. The primary evaluated outcome was vaccine uptake.

**Results:** Of the 1,058 participants who completed the survey, 704 (66.5%) were female, and 626 (59.2%) were nurses. Of all the respondents, 352 (33.27%) were enrolled to receive or had already received the vaccine, while 706 (66.73%) had not registered. In a bivariate analysis, not enrolling for vaccination was more likely in females than males (78.5% vs. 21.5%, P < 0.001), HCWs between the ages of 20 and 40 years than those > 40 years (70.4% vs. 29.6%, P = 0.005), Saudi HCWs than expatriates (78% vs 22%, P < 0.001), and among HCWs who used social media as a source of information than those who did not (69.8% vs. 38.6%, P < 0.001). In a multivariate analysis, independent factors for not enrolling to receive the vaccine included being female (aOR = 0.287, 95%CI = 0.206–0.401, P < 0.001), being less than 40 years of age (aOR = 1.021, 95%CI = 1.002–1.040, P = 0.032), and using social media as a source of information (aOR = 0.207, 95%CI = 0.132-1.354, P = 0.001). Factors associated with uptake were being a Saudi national (aOR = 1.918, 95%CI = 1.363–2.698, P < 0.001), working in an intensive care unit (aOR = 1.495, 95%CI = 1.083–2.063, P = 0.014), and working at a university hospital (aOR = 1.867, 95%CI = 1.380–2.525, P < 0.001).

**Conclusions:** A low level of vaccine uptake was observed especially in female HCWs, those younger than 40 years old, and those who used social media as their source of vaccine information. This survey provides important information for public health authorities in order to scale up vaccination campaigns targeting these HCWs to increase vaccine enrollment and uptake.

## 1 Introduction

After the coronavirus disease 2019 (COVID-19) reached pandemic levels, vaccine development was fast tracked through government funding, corporate spending, and private donations [1]. Once vaccines were made available in December 2020, a phased approach for vaccine allocation was recommended, with Phase 1a targeting first respondents and healthcare workers (HCWs) [2]. Several vaccine manufacturers have published their Phase 3 trials confirming the safety and efficacy of the vaccine [3-5]. However, such unprecedented scientific achievement is challenged by the hesitancy of HCWs to accept vaccination [6]. In an earlier study from the Kingdom of Saudi Arabia (KSA), 70% of the 1521 HCWs surveyed were willing to receive the COVID-19 vaccine [6]. Another study showed that 63% of the nurses surveyed were willing to receive the COVID-19 vaccine [7]. In two studies, the acceptance of the COVID-19 vaccine among adults was found to be between 58% and 69% [8, 9]. The KSA granted Pfizer/BioNTech emergency use authorization for the BNT162b2 vaccine on December 10, 2020, becoming the fourth country to do so after the United Kingdom, Bahrain, and Canada [10, 11]. On that same day, the Ministry of Health (MoH) sent out mass short message service texts and emails to all HCWs in the country encouraging them to voluntarily enroll for vaccine uptake through a dedicated smartphone application or the MoH website. COVID-19 vaccine rollout began on December 17, 2020. This study was conducted to evaluate vaccine enrollment and uptake within the first month of its rollout among HCWs in the KSA.

## 2 Method

### 2.1 Data collection

This national cross-sectional survey was conducted among HCWs in Saudi Arabia during the COVID-19 pandemic. Data were collected between December 27, 2020 and January 3, 2021. At the time of data collection, the national coronavirus vaccination campaign had already begun in the KSA, with HCWs as one of the prioritized groups. HCWs were surveyed regarding their intention to receive the COVID-19 vaccine. Participants were invited using a convenience sampling technique. We used several social media platforms and email lists to recruit participants. The survey was a pilot-validated, self-administered questionnaire that was sent to HCWs through SurveyMonkey^©^, a platform that allows researchers to deploy and analyze surveys via the web. The questionnaire was adapted from our previously published studies with modification and additions related to the new severe acute respiratory syndrome coronavirus 2 (SARS-CoV-2) variant of concern (VoC) [6, 12, 13].

The questions asked about respondents’ demographic characteristics (job category, age, gender, years of clinical experience, and work area), previous exposure to COVID-19 patients, previous COVID-19 infection, and travel history in the prior 3 months. We assessed the level of intention to and actual receipt (i.e., uptake) of the COVID-19 vaccine among HCWs. In addition, we assessed factors affecting respondents’ intention to receive the COVID-19 vaccine, including their level of awareness of the new SARS-CoV-2 VoC and sources of information. HCWs’ anxiety was measured by the validated 7-item General Anxiety Disorder (GAD-7) questionnaire, which has been used in several studies assessing HCWs’ anxiety levels during the pandemic [13, 14].

HCWs were informed of the purpose of the study in English at the beginning of the online survey. The respondents were given the opportunity to ask questions via a dedicated email address for the study. The Institutional Review Board at the College of Medicine and King Saud University Medical City approved the study (approval #20/0065/IRB). A waiver for signed consent was obtained since the survey presented no more than a minimal risk to subjects and involved no procedures for which written consent is usually required. To maximize confidentiality, personal identifiers were not required.

### 2.2 Statistical analyses

Descriptive analyses with means and standard deviations were applied to continuous variables, and categorically measured variables were described with frequencies and percentages. Histograms and statistical Kolmogorov–Smirnov tests of normality were used to assess the statistical normality of continuous variables. HCWs’ awareness of the new mutagenic COVID-19 virus strain was measured with eight questions, which received a score of 1 for each correctly answered knowledge/awareness question and 0 for each incorrectly answered question. Total awareness of the mutagenic viral outbreak was measured by adding up the total scores on the knowledge indicators, yielding a mutagenic disease awareness ranging from 0 to 8 points.

Independent samples t-tests were used to assess the statistical significance of mean scores between the levels of dichotomous categorical variables. Chi-squared tests of independence were used to assess the associations between categorically measured variables with the HCWs’ uptake of the COVID-19 vaccine. The logistic binary regression analysis was used to understand HCWs’ immunization uptake by regressing their sociodemographic, clinical, and professional characteristics and mutant viral strain perceptions against their odds of having actively received the COVID-19 immunization shot or registering for it. The associations between HCWs’ measured independent variables and COVID-19 vaccine uptake were expressed as adjusted odds ratios (aOR) with 95% confidence intervals (95%CI). IBM® SPSS® was used for the statistical data analysis, and significance was considered at the 0.05 alpha level.

## 3 Results

Of the 1,212 HCWs who accessed the survey, 1,058 (87.2%) completed the survey. Their sociodemographic characteristics are shown in Table 1.

**Table 1:** Descriptive analysis of HCWs’ sociodemographic and professional characteristics (N = 1058)

| Characteristic | n | % |
| --- | --- | --- |
| **Gender** |  |  |
| Male | 354 | 33.5 |
| Female | 704 | 66.5 |
| **Age** |  |  |
| 20–30 years | 238 | 22.5 |
| 31–40 years | 471 | 44.5 |
| 41–50 years | 263 | 24.9 |
| ≥ 51 years | 86 | 8.1 |
| **Marital state** |  |  |
| Single | 257 | 24.3 |
| Married/divorced/widowed | 801 | 75.7 |
| **Nationality** |  |  |
| Expatriate | 736 | 69.6 |
| Saudi | 322 | 30.4 |
| **Clinical Role** |  |  |
| Consultant | 213 | 20.1 |
| Assistant consultant/fellow | 52 | 4.9 |
| Resident/registrar | 138 | 13.0 |
| Nurse | 626 | 59.2 |
| Intern/medical student | 29 | 2.7 |
| **Hospital Area** |  |  |
| ICU | 273 | 25.8 |
| ER | 110 | 10.4 |
| OR | 62 | 5.9 |
| Isolation ward | 63 | 6.0 |
| General ward | 261 | 24.7 |
| OPD | 225 | 21.3 |
| Non-clinical area | 64 | 6.0 |
| **Hospital sector** |  |  |
| Private | 174 | 16.4 |
| Public/governmental | 487 | 46.0 |
| University hospital | 397 | 37.5 |
| **Hospital specialty** |  |  |
| Primary healthcare center | 123 | 11.6 |
| Secondary care hospital | 196 | 18.5 |
| Tertiary hospital | 739 | 69.8 |

ICU: intensive care unit; ER: emergency room; OR: operating room; OPD: outpatient department.

Of all the respondents, 352 (33.27%) were enrolled to receive or had already received the vaccine, while 706 (66.73%) did not wish to register for vaccination. The bivariate analysis of association between the respondents’ characteristics and their tendency to receive the vaccine is shown in Table 2. A significantly higher percentage of females compared to males reported not receiving or registering to receive the vaccine (78.5% vs. 21.5%, P < 0.001), and younger age (between 20 and 40 years old) was associated with a significant tendency to decline to receive the vaccine compared to older age groups (P = 0.005). A lower percentage of expatriates reported receiving or registering to receive the vaccine compared to Saudi nationals (P < 0.001).

**Table 2:**
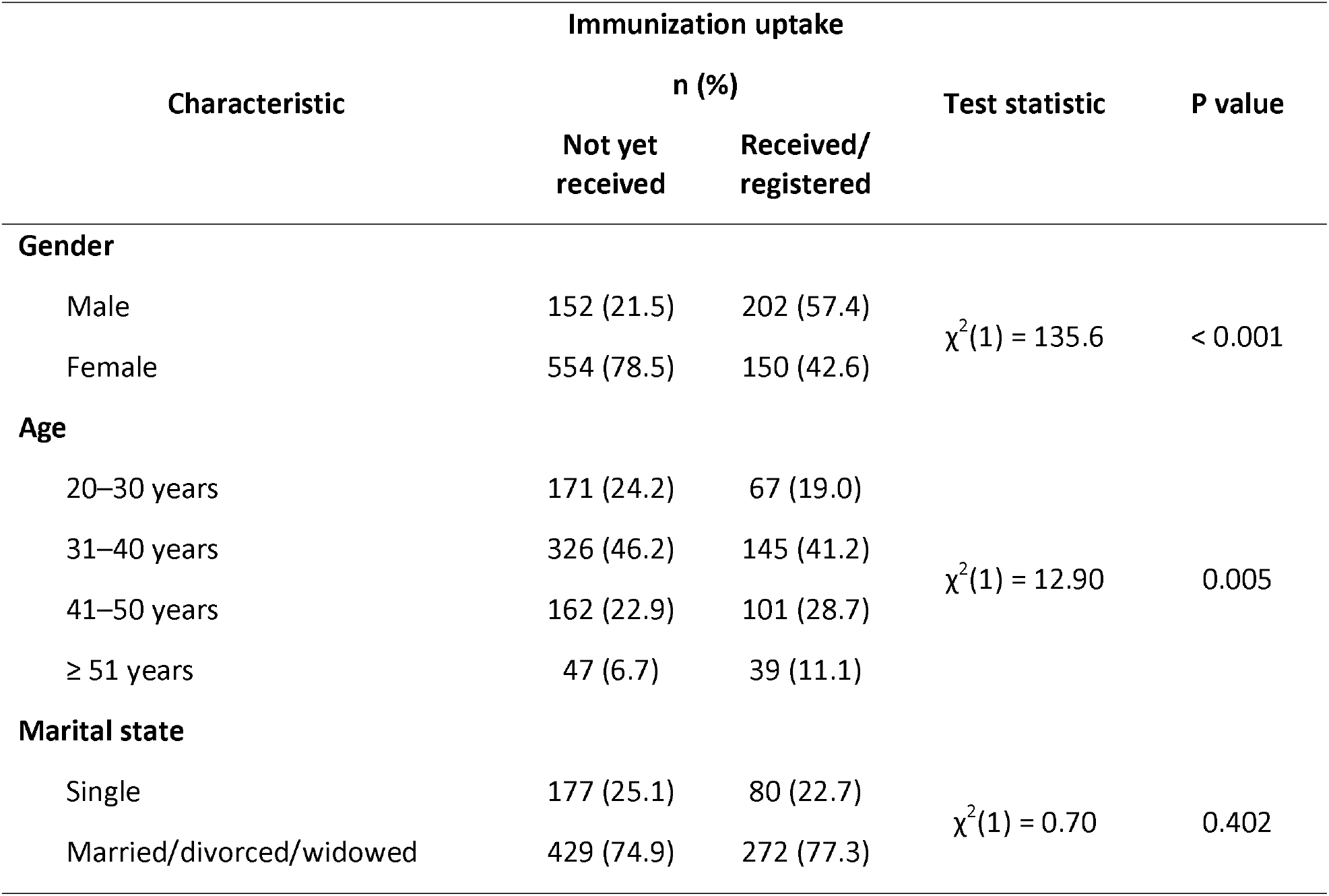
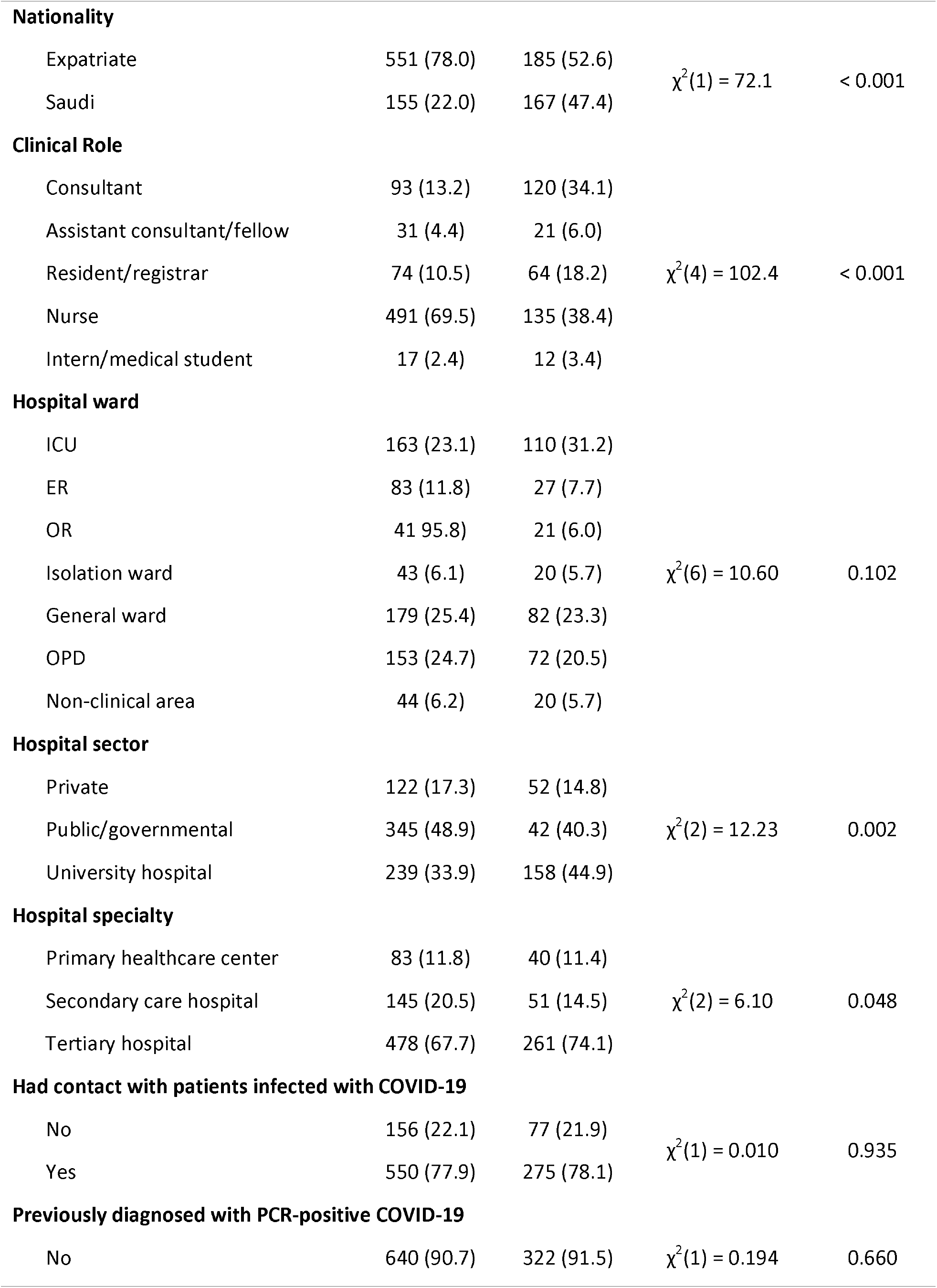
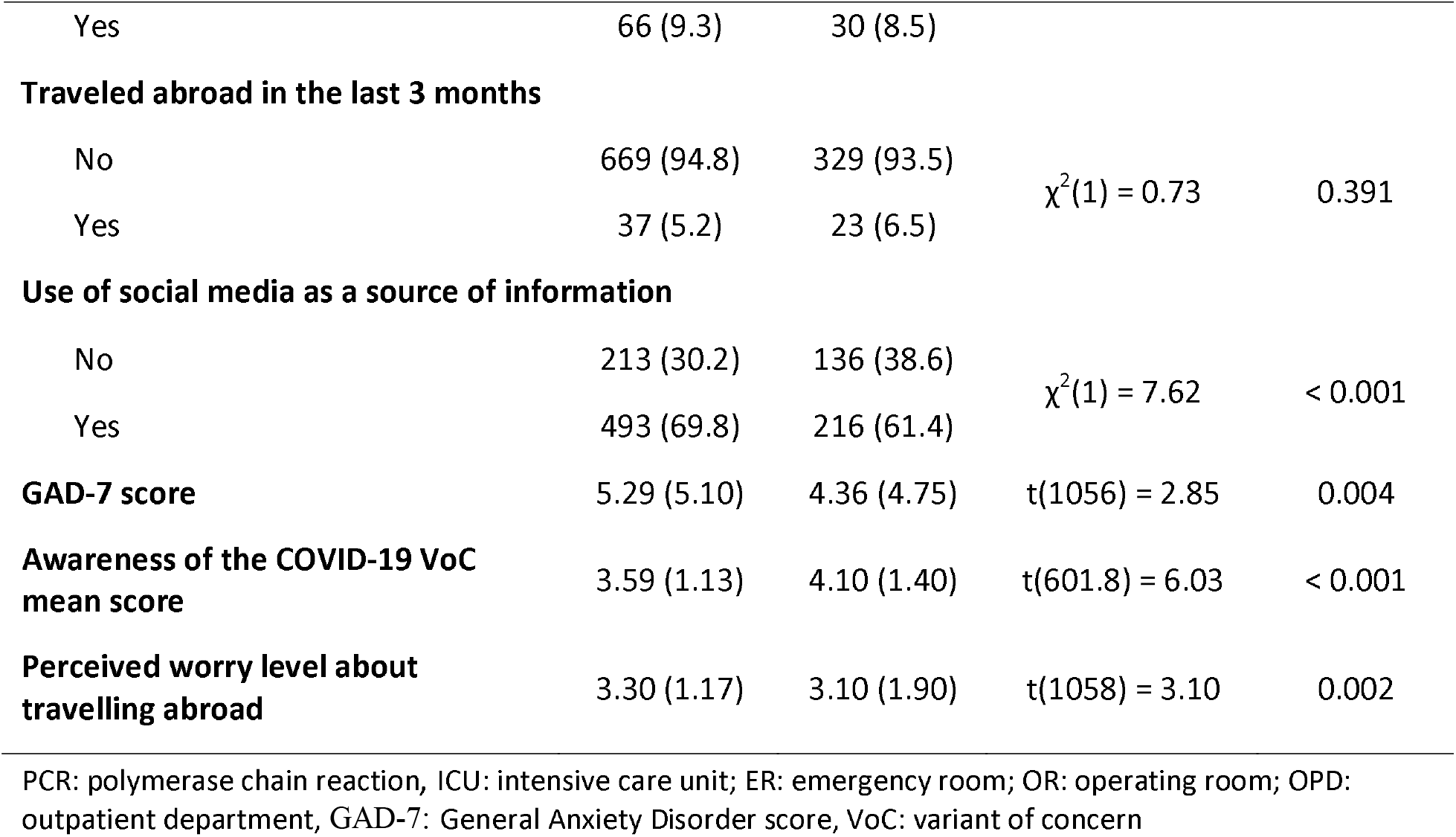
Descriptive bivariate analysis of HCWs’ uptake of the COVID-19 vaccine (N = 1058)

| Characteristic | Immunization uptake |  | Test statistic | P value |
| --- | --- | --- | --- | --- |
|  | n (%) |  |  |  |
|  | Not yet received | Received/registered |  |  |
| **Gender** |  |  |  |  |
| Male | 152 (21.5) | 202 (57.4) | $\chi^2(1) = 135.6$ | < 0.001 |
| Female | 554 (78.5) | 150 (42.6) |  |  |
| **Age** |  |  |  |  |
| 20–30 years | 171 (24.2) | 67 (19.0) | $\chi^2(1) = 12.90$ | 0.005 |
| 31–40 years | 326 (46.2) | 145 (41.2) |  |  |
| 41–50 years | 162 (22.9) | 101 (28.7) |  |  |
| ≥ 51 years | 47 (6.7) | 39 (11.1) |  |  |
| **Marital state** |  |  |  |  |
| Single | 177 (25.1) | 80 (22.7) | $\chi^2(1) = 0.70$ | 0.402 |
| Married/divorced/widowed | 429 (74.9) | 272 (77.3) |  |  |

|  |  |  |  |  |  |
| --- | --- | --- | --- | --- | --- |
| Nationality |  |  |  |  |  |
| Expatriate | 551 (78.0) | 185 (52.6) | $\chi^2(1) = 72.1$ | < 0.001 | |
| Saudi | 155 (22.0) | 167 (47.4) |  |  |  |
| Clinical Role |  |  |  |  |  |
| Consultant | 93 (13.2) | 120 (34.1) | $\chi^2(4) = 102.4$ | < 0.001 | |
| Assistant consultant/fellow | 31 (4.4) | 21 (6.0) |  |  |  |
| Resident/registrar | 74 (10.5) | 64 (18.2) |  |  |  |
| Nurse | 491 (69.5) | 135 (38.4) |  |  |  |
| Intern/medical student | 17 (2.4) | 12 (3.4) |  |  |  |
| Hospital ward |  |  |  |  |  |
| ICU | 163 (23.1) | 110 (31.2) | $\chi^2(6) = 10.60$ | 0.102 | |
| ER | 83 (11.8) | 27 (7.7) |  |  |  |
| OR | 41 95.8) | 21 (6.0) |  |  |  |
| Isolation ward | 43 (6.1) | 20 (5.7) |  |  |  |
| General ward | 179 (25.4) | 82 (23.3) |  |  |  |
| OPD | 153 (24.7) | 72 (20.5) |  |  |  |
| Non-clinical area | 44 (6.2) | 20 (5.7) |  |  |  |
| Hospital sector |  |  |  |  |  |
| Private | 122 (17.3) | 52 (14.8) | $\chi^2(2) = 12.23$ | 0.002 | |
| Public/governmental | 345 (48.9) | 42 (40.3) |  |  |  |
| University hospital | 239 (33.9) | 158 (44.9) |  |  |  |
| Hospital specialty |  |  |  |  |  |
| Primary healthcare center | 83 (11.8) | 40 (11.4) | $\chi^2(2) = 6.10$ | 0.048 | |
| Secondary care hospital | 145 (20.5) | 51 (14.5) |  |  |  |
| Tertiary hospital | 478 (67.7) | 261 (74.1) |  |  |  |
| Had contact with patients infected with COVID-19 |  |  |  |  |  |
| No | 156 (22.1) | 77 (21.9) | $\chi^2(1) = 0.010$ | 0.935 | |
| Yes | 550 (77.9) | 275 (78.1) |  |  |  |
| Previously diagnosed with PCR-positive COVID-19 |  |  |  |  |  |
| No | 640 (90.7) | 322 (91.5) | $\chi^2(1) = 0.194$ | 0.660 | |
| Yes | 66 (9.3) | 30 (8.5) |  |  |
| Traveled abroad in the last 3 months |  |  |  |  |
| No | 669 (94.8) | 329 (93.5) | $\chi^2(1) = 0.73$ | 0.391 |
| Yes | 37 (5.2) | 23 (6.5) |  |  |
| Use of social media as a source of information |  |  |  |  |
| No | 213 (30.2) | 136 (38.6) | $\chi^2(1) = 7.62$ | < 0.001 |
| Yes | 493 (69.8) | 216 (61.4) |  |  |
| GAD-7 score | 5.29 (5.10) | 4.36 (4.75) | t(1056) = 2.85 | 0.004 |
| Awareness of the COVID-19 VoC mean score | 3.59 (1.13) | 4.10 (1.40) | t(601.8) = 6.03 | < 0.001 |
| Perceived worry level about travelling abroad | 3.30 (1.17) | 3.10 (1.90) | t(1058) = 3.10 | 0.002 |

PCR: polymerase chain reaction, ICU: intensive care unit; ER: emergency room; OR: operating room; OPD: outpatient department, GAD-7: General Anxiety Disorder score, VoC: variant of concern

A significantly higher percentage of nurses (69.5%) and HCWs working in public/governmental hospitals (49.8%) had not received or registered to receive the vaccine in comparison to other clinical roles (P < 0.001) and HCWs in other hospital sectors (P = 0.002). HCWs working in university hospitals (44.9%, P = 0.002) and tertiary care hospitals (74.1%, P = 0.048) were more inclined to receive the vaccine.

HCWs’ previous infection with laboratory-confirmed COVID-19, previous contact with COVID-19 patients, and their travel history over the last 3 months were not correlated with their vaccine uptake. The respondents who were inclined to receive the vaccine were significantly less dependent on using social media as a source of information and had a significantly lower GAD-7 score, higher awareness about the new VoC, and lower level of worry about travelling abroad.

A binomial logistic regression was performed to analyze the independent association between HCWs’ characteristics and their vaccine uptake behavior as shown in Table 3. Females were significantly less likely to receive or register to receive the vaccine (aOR = 0.287, P < 0.001), while older age (aOR = 1.021, P = 0.032) and Saudi nationality (aOR = 1.918, P = 0.001) were associated with an increased likelihood of vaccine uptake. Intensive care unit (ICU) staff (aOR = 1.495, P = 0.014) and staff working in university hospitals (aOR = 1.867, P < 0.001) were also significantly and independently more likely to receive or register to receive the vaccine. A higher level of awareness of the VoC also significantly predicted vaccine uptake among HCWs (aOR = 1.131, P = 0.047). HCWs’ level of anxiety as measured by their GAD-7 score, history of travelling abroad over the previous 3 months, and personal history of previous polymerase chain reaction (PCR)-positive COVID-19 did not independently predict their vaccine uptake behavior. Figure 1 illustrates the linear incremental relation between HCWs’ age and the probability of vaccine uptake, as the probability rose from about 30% for the 20–31 age group to almost 45% for those over 50 years of age.

**Table 3:** Multivariate logistic binary regression analysis of HCWs’ COVID-19 immunization behavior (registering or receiving the vaccine) (N = 1058)

| Characteristic | Multivariate aOR | 95%CI |  | P value |
| --- | --- | --- | --- | --- |
|  |  | Lower | Upper |  |
| Gender = Female | 0.287 | 0.206 | 0.401 | < 0.001 |
| Age above 40 years | 1.021 | 1.002 | 1.040 | 0.032 |
| Marital status =<br>Married/divorced/widowed | 0.799 | 0.545 | 1.169 | 0.247 |
| Nationality = Saudi | 1.918 | 1.363 | 2.698 | < 0.001 |
| Hospital ward = ICU | 1.495 | 1.083 | 2.063 | 0.014 |
| Hospital sector = University<br>hospital | 1.867 | 1.380 | 2.525 | < 0.001 |
| Mean score awareness of<br>variant of concern (range: 0–8<br>points) | 1.131 | 1.002 | 1.278 | 0.047 |
| GAD-7 score | 0.995 | 0.966 | 1.025 | 0.742 |
| Travelled abroad in last 3<br>months | 1.624 | 0.889 | 2.964 | 0.114 |
| Previously diagnosed with<br>COVID-19 | 0.880 | 0.536 | 1.445 | 0.614 |
| Use of social media as a source<br>of information | 0.207 | 0.132 | 1.354 | 0.001 |

ICU: intensive care unit, GAD-7: General Anxiety Disorder score

**Figure 1:**
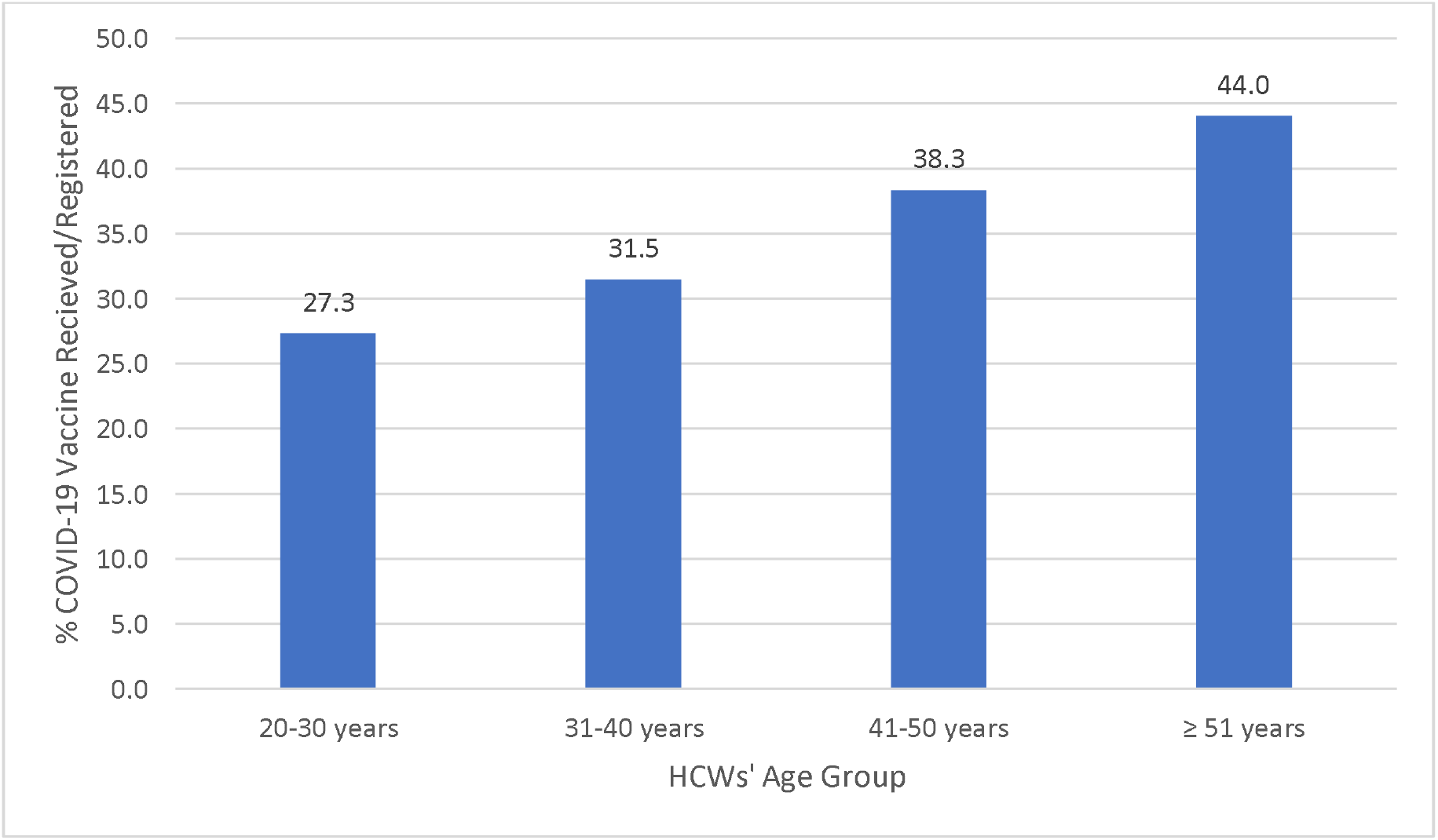
The association between HCWs’ age group and their model mean predicted probability of COVID-19 vaccination.

## 4 Discussion

In this reported national survey on COVID-19 vaccine uptake in one of the first countries to roll out the BNT162b2 vaccine, only 352 (33.3%) of 1,058 HCWs had either registered to/received the vaccine within 3 weeks of its availability. In a previous cross-sectional survey to assess HCWs’ COVID-19 vaccine confidence and hesitancy prior to launching a vaccine campaign in the KSA, 70% were willing to receive a vaccine once available[6]. Additionally, half of the participants indicated that they would receive the vaccine as soon as it became available, while more than one-third preferred delaying receiving it for a few months. In a study that specifically focused on vaccine acceptance according to vaccine type, only 20.9% were willing to receive BNT162b2 [15]. The low vaccine uptake reported in the current study, together with HCWs’ earlier reports of preferring to delay getting vaccinated, is alarming and should trigger public health officials to target these groups with campaigns to enhance their vaccine confidence and acceptance.

In the current study, two-thirds of participants were female, almost 60% were nurses, 70% were expatriates, and the majority worked at tertiary care hospitals. These findings are similar to a previous study that was conducted prior to the vaccine rollout [6]. However, that study included only 50% nurses. In this study, 69% of nurses had neither received nor registered to receive the vaccine, while half of the physicians had. This is similar to influenza vaccine uptake among HCWs, as it has been reported that physicians have significantly higher flu vaccination rates compared to nurses [16, 17].

Almost all of the participants worked in clinical areas, and 80% managed COVID-19 patients. HCWs working in clinical areas other than the ICU, such as the emergency room (ER) and wards, did not converge significantly on their vaccine uptake. In an influenza vaccine uptake study, only working in the building where the vaccination was being performed made a significant difference [18].

HCWs from university hospitals were found to be significantly more likely to receive the vaccine than those working in private and public sectors. Additionally, HCWs in tertiary healthcare settings were significantly more likely to get the vaccine than those working in primary and secondary healthcare settings. In a systematic review on influenza vaccination among HCWs, the top reason for vaccine uptake was self-protection rather than protecting patients or setting an example for their patients, with no observed difference in hospital settings [19].

A gender difference was observed in the vaccine uptake, with female HCWs being significantly less likely to receive the vaccine than male HCWs (P < 0.001). The discrepancy between males and females in the uptake of this vaccine is interesting. Male sex was shown to be associated with increased death and ICU admission in a recent meta-analysis of COVID-19 patients [20]. However, anti-Spike antibodies declined faster in female than male patients in a recent French study [21]. These differences are important to further enhance our understanding of vaccine uptake and design-specific interventions.

While another study showed no effect of age on COVID-19 vaccine acceptance among the general population [22], our study revealed that HCWs over 40 years of age were more motivated to receive the vaccine. This is in contrast to a vaccine intent survey among nurses that showed a stronger COVID-19 vaccination intention among younger workers [7]. While older HCWs are at a higher risk of COVID-19 infection, protection of the entire healthcare workforce is crucial during this pandemic.

Saudi HCWs were found to be significantly more likely to receive the vaccine than expatriates (P < 0.001). The KSA has made the COVID-19 vaccine available free of charge to all citizens and residents. A previous study found a disparity in the outcome of patients infected with SARS-CoV-2 in relation to gender and ethnicity [23]. In a study among blood donors in the KSA, non-Saudis were found to be more likely to have positive SARS-CoV-2 serology [24]. These differences between Saudi and expatriates deserve further study in order to understand the factors contributing to this disparity, which could allow for strategies and communication plans to alleviate the risk of exposure to SARS-CoV-2 and enhance the acceptance of the COVID-19 vaccine among the population.

HCWs’ clinical role was correlated with their uptake of the vaccine in the bivariate analysis. Nurses were found to be significantly less inclined to receive the vaccine than physicians and other professionals (P < 0.001), which is similar to a previous study from the KSA [6]. The multivariate analysis did not show any significant differences between location with the exception of ICU staff, who were significantly more inclined to receive the vaccine (aOR = 1.495, P = 0.014). HCWs are more likely to acquire vaccine-preventable diseases, with 20% of HCWs contracting influenza annually, recent reports showed low influenza vaccine uptake among doctors and nurses (56.5% and 34.8% acceptance rate, respectively)[25].

Interestingly, no significant differences in vaccine uptake were found between HCWs who managed COVID-19 patients compared to those who did not or between HCWs with previous COVID-19 infections compared to those without. The low COVID-19 vaccine uptake rate in the middle of a pandemic is alarming, and efforts should focus on increasing vaccine acceptance and uptake to match the speed of the pandemic.

The VoC-202012/01 emerged in December 2020, resulting in new travel restrictions [26]. However, there is evidence that the BNT162b2 vaccine is effective against this variant based on in-vitro studies [27]. In this study, vaccine uptake was not influenced by HCWs’ travel history. However, the overall sample size of returning travelers was small and may not be representative. Information on evolving variants are emerging in various countries [28], and concerns regarding the vaccine’s efficacy against these variants may hinder vaccine uptake. This is a concerning situation that warrants further study.

Even before the COVID-19 pandemic, vaccination was considered an emotionally charged topic in many cultures [29]. However, vaccine hesitancy is common and includes people who have not yet rejected vaccination but do not trust the institutions connected to the vaccine [30]. Current recommendations suggest not only to make a safe and effective vaccine available but also deep engagement of around the human element to build public trust in any vaccine [31]. This highlights the importance of addressing societal concerns and fears to ensure a vaccination campaign’s success [32]. Personal worries and baseline anxiety should not be neglected as these could trigger vaccine refusal in the community via the butterfly effect. Findings in the current study highlight how HCWs, especially those with lower GAD-7 scores, were more likely to accept the new vaccine. It also provides a glimpse of the relationship between higher awareness (in this case, of the new variant) and the likelihood of considering vaccination.

The use of social media for information could greatly affect HCWs’ and the general populations’ COVID-19 vaccine acceptance. While some studies did not find associations between willingness to vaccinate and social media use [22, 33], others found a higher vaccination willingness among respondents from the general population who did not rely on social media for COVID-19 information [34, 35]. One study assessed the attitudes towards COVID-19 vaccines using the Vaccine Conspiracy Belief Scale and showed higher conspiracy beliefs among respondents who relied on social media platforms as their main source of information [35].

### 4.1 Study limitations and strengths/future potential

This study is subject to the limitations of cross-sectional surveys, including sampling, response, and recall biases. While this work did not explore the reasons why HCWs did not register for the vaccine, it presents their initial vaccination acceptance, which needs to be explored in future studies. It should be noted that HCWs’ perceptions and vaccine hesitancy may differ from one country to another.

### 4.2 Conclusion

This study observed a low level of COVID-19 vaccine enrollment among HCWs during the first month of the vaccine rollout in one of the first countries to roll out the vaccine. Public health officials should scale up their efforts to increase vaccine acceptance and uptake among HCWs to match the speed of the growing pandemic. Optimizing protection of HCWs through vaccination and encouraging them to subsequently recommend vaccination to their patients is vital to curbing this global crisis.

## Data Availability

All the data for this study will be made available upon reasonable request.

## Ethics approval and consent to participate

The study was approved by the institutional review board of King Saud University (approval #20/0065/IRB).

